# Are children with disabilities more likely to be malnourished than children without disabilities? Evidence from the Multiple Indicator Cluster Surveys in 30 countries

**DOI:** 10.1101/2023.09.25.23296066

**Authors:** Sara Rotenberg, Shanquan Chen, Xanthe Hunt, Tracey Smythe, Hannah Kuper

## Abstract

**Introduction:** A key Sustainable Development Goal target is to eliminate all forms of malnutrition. Existing evidence suggests children with disabilities are at greater risks of malnutrition, exclusion from nutrition programmes, and mortality from severe acute malnutrition than children without disabilities. However, there is limited evidence on the nutritional outcomes of children with disabilities in large-scale global health surveys.

**Methods:** We analysed Multiple Indicator Cluster Survey (MICS) data from 30 low and middle-income countries (LMICs) to compare nutritional outcomes for children aged 2-4 years with and without disabilities. We estimated the adjusted prevalence ratios for stunting, wasting, and underweight comparing children with and without disabilities by country and sex, using quasi-Poisson models with robust standard errors. We accounted for the complex survey design, wealth quintile, location, and age in the analyses. We meta-analysed these results to create an overall estimate for each of these outcomes.

**Results:** Our analyses included 229,621 children aged 2-4 across 30 countries, including 15,071 children with disabilities (6.6%). Overall, children with disabilities were more likely to be stunted (aRR: 1.16, 95% C.I.: 1.11 -1.20), wasted (aRR: 1.28, 95% C.I.: 1.18 – 1.39), and underweight (aRR: 1.33, 95% C.I.: 1.17, 1.51) than children without disabilities. These patterns were observed in both girls and boys with disabilities, compared to those without.

**Conclusion:** Children with disabilities are significantly more likely to experience all forms of malnutrition, making it critical to accelerate efforts to improve disability-inclusion within nutrition programmes. Ending all forms of malnutrition will not be achievable without a focus on disability.

**Key Messages:** *What is already known on this topic:*

- Prior research has shown children with disabilities in low-and middle-income countries have higher prevalence of stunting, wasting, and underweight and worse outcomes and mortality from severe acute malnutrition.

*What this study adds:*

- We show that children with disabilities, overall and by sex, have significantly higher rates of stunting, wasting, and underweight than children without disabilities.
- This study adds to the existing evidence on disability-based inequities in nutritional outcomes from nationally representative, internationally-comparable household surveys in multiple countries.

*How this study might affect research, practice, or policy:*

- A twin-track approach is needed to ensure children with disabilities are reached in mainstream nutrition programmes, as well as having their specific and additional needs met through targeted programmes.
- Without sufficient focus on disability, it will be impossible to achieve SDG2, to end all forms of child nutrition, or meet global child mortality reduction targets.

## Introduction

Malnutrition is a major contributor to child mortality worldwide.^1^ It often arises from a complex interaction of factors, including socioeconomic status, gender inequality, political instability, food insecurity, and poor nutritional intake.^2^ However, access to and experiences of adequate nutrition vary among children, and challenges with these can hinder their development and compromise their well-being. Certain groups, such as children with disabilities, may be at particular risk of inadequate nutrition. Prior research has shown that children with disabilities have higher prevalence of malnutrition and its sequelae. This is a consequential relationship for the nearly 240 million children with disabilities worldwide.^3^ For example, a 2017 systematic review found that children with disabilities had nearly three times higher odds of being underweight (OR 2.97, 95% C.I.: 2.33 - 3.79) and two times higher odds of being stunted or wasted (Stunting: OR: 1.82, 95% C.I. 1.40 - 2.36; Wasting: OR:1.90, 95% C.I.: 1.32 - 2.75) compared to children without disabilities.^4^ However, these studies used variable definitions of disability and malnutrition, making international comparison difficult. A longitudinal cohort study in Malawi showed that children with disabilities also have significantly higher mortality rates from severe acute malnutrition than children without disabilities (mortality HR: 2.29, 95%CI: 1.51 – 3.45).^5^ Further, while some types of impairments may make the use of standardised measures of nutritional status invalid (e.g., growth restriction or limb difference),^6^ these conditions do not occur at sufficiently high prevalence to distort estimates drawn from large samples. Indeed, previous descriptive analysis of the MICS has shown that children with functional difficulty in the walking, playing, and fine-motor domains have the highest prevalence of stunting, wasting, and underweight.^3^

It is well-established that the relationship between impairment and malnutrition is likely to be bidirectional,^7^ with children with disabilities more at risk of malnutrition^4^ and children with severe acute malnutrition more at risk of acquiring impairments.^8–10^ Some proportion of the difference may be linked to a child’s impairment. For example, there is evidence that functional limitations, feeding difficulties, and inadequate energy intake are key risk factors that lead children with cerebral palsy to be malnourished.^11^ While nutritional disorders are common among some impairment types (such as cerebral palsy) ^11–14^ these inequities are inexplicable by impairment alone. Moreover, several of the social factors that lead to worse nutritional outcomes are also more prevalent in children with disabilities. For example, inequities in maternal education, poverty, parental employment status, and access to water, sanitation and hygiene (WASH) and information and communication technology (ICT) are closely linked to inequities in both nutritional status^15^ and disability.^16–20^ Similarly, recent research has highlighted that children with disabilities have higher occurrence of common childhood illnesses, such as acute respiratory infection, fever, and diarrhoeal disease,^21^ ^22^ which are known to co-occur with wasting and other equity-related variables.^23^

Despite the evidence for this bidirectional relationship, as well as the overlap between regions with high malnutrition prevalence^24^ and those with high childhood disability prevalence,^25^ disability is not sufficiently attended to in guidelines on malnutrition, putting children with disabilities at greater risk of adverse outcomes from malnutrition and other nutritional disorders.^26^ Since tackling all forms of malnutrition is one of the targets of Sustainable Development Goal 2, it is also important to understand how children with disabilities are being reached in these efforts.^27^ Without a focus on disability, there is the risk of leaving these children behind.^28^ This is likely to require a twin-track approach, which involves simultaneously addressing the specific needs and challenges faced by a particular group, such as children with disabilities, while also implementing broader strategies to achieve a larger goal, such as improving nutritional status and addressing malnutrition for all children. However, more evidence is needed on the association between disability and nutritional status.

The Multiple Indicator Cluster Survey (MICS) provides an opportunity to fill the evidence gap by drawing on internationally-comparable data with comparable measures of disability and malnutrition. While a recent UNICEF report presented some descriptive analysis for all countries combined and by impairment,^3^ this analysis will look at relative and absolute inequities across gender and disability. The aim of this paper is therefore to use MICS data to examine relative inequities in malnutrition indicators by disability status and sets out to answer the question: are children with disabilities more likely to be stunted, wasted, or underweight than children without disabilities?

## Methods

### Data source

We used data from the sixth round of the UNICEF-supported Multiple Indicator Cluster Survey conducted between 2017-2021 in 30 countries. All data were publicly available on the MICS data repository as of April 2023.^29^ The MICS utilize a multi-stage probability sampling methodology to generate nationally representative data on indicators for monitoring progress towards the Sustainable Development Goals, health, and human development.^30^ The current analyses focus on MICS data from 30 countries where information was available on both disability status and nutrition among children aged 2-4. Trained interviewers conducted household-based surveys with randomly selected households. All children aged 2-4 within selected households were eligible to participate.^30^ The survey questions were standardized across countries to enable comparative analyses. We included data from all publicly available MICS surveys as of April 2023 that contained information on the variables of interest.

#### Exposure

Disability was measured using the child functioning module for children aged 2-4 years old. Caregivers were asked about their child’s functioning across eight functional domains: vision, hearing, communication, walking, controlling behaviour, learning, fine motor skills, and playing. Children were considered disabled if their caregiver reported ‘a lot of difficulty’ or ‘cannot do at all’ in at least one functional domain.

#### Outcomes

Data available for download on the MICS data are cleaned to provide a z-score for children’s weight for age (underweight), weight for height (wasting), and height for age (stunting) compared to the WHO Child Growth Standards.^31^ Children whose standardized z-score are 2 or more standard deviations from the WHO Child Growth Standards are recoded as underweight, wasted, or stunted.^32^

#### Covariates

Age was reported by caregivers, while location was determined according to the area in which participants were selected for the survey. Wealth status was calculated by UNICEF according to data on household characteristics, household and personal assets, and WASH via principal components analysis.^33^

### Statistical Analysis

All analyses were conducted using R statistical software version 4.2.2 (R Core Team, 2022) and statistical significance was determined as p < 0.05. Outcomes, exposures, and covariates were described by country and sex using summary statistics. Continuous data were reported as mean (standard deviation [SD]) and categorical data as frequencies (percentage).

To estimate the relative inequality in each outcome between children with and without disabilities, modified Poisson regression models were fitted to estimate the risk ratio (RR)^34^ and 95% confidence interval (CI) for each outcome by country and by country and sex, adjusting for age, residence place, and wealth status. The complex survey design and sample weights were accounted for using the ’survey’ package in R.^35^ Country-specific RRs were pooled via random-effects meta-analysis if significant heterogeneity was detected across countries per Cochran’s Q test (p <0.1), otherwise fixed-effects meta-analysis was used.

Records with missing data were excluded from analyses rather than imputed. To minimize bias from small sample sizes, countries with fewer than 25 respondents with disabilities were excluded when pooling estimates. This secondary analysis of anonymized data was approved by the London School of Hygiene and Tropical Medicine Research Ethics Committee on November 9, 2020 (Ref: 22719).

## Results

220,621 children aged 2-4 were eligible for inclusion across 30 countries (Table 1). Country sample sizes ranged from 1,268 children in Kiribati to 67,612 children in Pakistan (including only Balochistan, Khyber Paktunkhwa, Sindh, and Punjab Provinces). The sample includes 15,071 children with disabilities (6.6%) overall, though country prevalence ranged from 2.0% (n=68) in Cuba to 14.4% (n=784) in Central African Republic. The sample had a mean age of 3.01 years (SD: 0.81) and was 51% male (n=117,132). Most of the sample lived in rural areas (67.6%, n=155,120). In the overall sample, 31.6% of children were stunted (n=72,489), 5.9% were wasted (n=13,606), and 18.6% were underweight (n=42,716).

**Table 1:** Baseline characteristics of the sample.

| Table 1: Baseline characteristics of the sample |  |  |  |  |  |  |  |  |  |
| --- | --- | --- | --- | --- | --- | --- | --- | --- | --- |
| Country | Total N | Disability (n, %) | Age (mean, SD) | Male (n, %) | Rural (n, %) | Wealth quintile (mean, SD) | Stunted (n, %) | Wasted (n, %) | Underweight (n, %) |
| Pooled | 229,621 | 15,071 (6.6) | 3.01 (0.81) | 117,132 (51.0) | 155,120 (67.6) | 2.66 (1.38) | 72,489 (31.6) | 13,606 (5.9) | 42,716 (18.6) |
| Algeria | 9,089 | 234 (2.6) | 3.01 (0.81) | 4,669 (51.4) | 3,426 (37.7) | 2.68 (1.37) | 885 (9.7) | 154 (1.7) | 217 (2.4) |
| Argentina | 3,991 | 148 (3.7) | 3.03 (0.82) | 2,074 (52.0) | 3,991 (100.0) | 2.74 (1.41) | 377 (9.4) | 80 (2.0) | 107 (2.7) |
| Bangladesh | 14,055 | 373 (2.7) | 3.00 (0.81) | 7,298 (51.9) | 11,471 (81.6) | 2.79 (1.42) | 4,078 (29.0) | 1,297 (9.2) | 3,439 (24.5) |
| Central African Republic | 5,436 | 784 (14.4) | 3.02 (0.81) | 2,697 (49.6) | 3,523 (64.8) | 2.97 (1.39) | 2,400 (44.2) | 215 (4.0) | 1,249 (23.0) |
| Chad | 14,155 | 1,524 (10.8) | 3.03 (0.81) | 7,185 (50.8) | 11,679 (82.5) | 2.76 (1.41) | 5,871 (41.5) | 2,006 (14.2) | 4,532 (32.0) |
| Costa Rica | 2,212 | 154 (7.0) | 2.96 (0.81) | 1,153 (52.1) | 920 (41.6) | 2.46 (1.37) | 141 (6.4) | 35 (1.6) | 61 (2.8) |
| Cuba | 3,356 | 68 (2.0) | 3.02 (0.79) | 1,687 (50.3) | 1,002 (29.9) | 3.04 (1.40) | 234 (7.0) | 92 (2.7) | 96 (2.9) |
| Dominican Republic | 5,075 | 201 (4.0) | 3.02 (0.82) | 2,538 (50.0) | 1,722 (33.9) | 2.51 (1.37) | 283 (5.6) | 98 (1.9) | 147 (2.9) |
| Democratic Republic of Congo | 12,742 | 943 (7.4) | 3.00 (0.81) | 6,286 (49.3) | 9,320 (73.1) | 2.35 (1.24) | 6,382 (50.1) | 716 (5.6) | 3,425 (26.9) |
| The Gambia | 6,166 | 417 (6.8) | 3.02 (0.80) | 3,153 (51.1) | 3,900 (63.3) | 2.34 (1.32) | 1,425 (23.1) | 364 (5.9) | 985 (16.0) |
| Ghana | 5,413 | 551 (10.2) | 3.01 (0.81) | 2,661 (49.2) | 3,263 (60.3) | 2.66 (1.44) | 1,041 (19.2) | 230 (4.2) | 602 (11.1) |
| Guinea Bissau | 4,602 | 188 (4.1) | 3.03 (0.82) | 2,363 (51.3) | 3,587 (77.9) | 2.51 (1.30) | 1,303 (28.3) | 202 (4.4) | 743 (16.1) |
| Honduras | 5,177 | 282 (5.4) | 3.03 (0.80) | 2,648 (51.1) | 3,415 (66.0) | 2.49 (1.34) | 1,184 (22.9) | 70 (1.4) | 412 (8.0) |
| Iraq | 10,179 | 341 (3.4) | 3.02 (0.80) | 5,198 (51.1) | 3,833 (37.7) | 2.64 (1.40) | 1,055 (10.4) | 177 (1.7) | 303 (3.0) |
| Kiribati | 1,268 | 156 (12.3) | 3.01 (0.82) | 634 (50.0) | 807 (63.6) | 2.55 (1.39) | 238 (18.8) | 20 (1.6) | 75 (5.9) |
| Lao | 7,178 | 220 (3.1) | 2.99 (0.81) | 3,617 (50.4) | 4,358 (60.7) | 2.61 (1.39) | 2,887 (40.2) | 572 (8.0) | 1,806 (25.2) |
| Lesotho | 2,027 | 169 (8.3) | 3.01 (0.83) | 987 (48.7) | 1,550 (76.5) | 2.48 (1.37) | 710 (35.0) | 33 (1.6) | 196 (9.7) |
| Madagascar | 7,626 | 750 (9.8) | 3.02 (0.81) | 3,855 (50.6) | 5,895 (77.3) | 2.44 (1.34) | 3,297 (43.2) | 457 (6.0) | 2,133 (28.0) |
| Malawi | 9,133 | 489 (5.4) | 2.97 (0.82) | 4,560 (49.9) | 8,041 (88.0) | 2.87 (1.39) | 3,213 (35.2) | 196 (2.1) | 1,172 (12.8) |
| Mongolia | 3,803 | 89 (2.3) | 3.02 (0.81) | 1,976 (52.0) | 1,882 (49.5) | 2.56 (1.35) | 432 (11.4) | 34 (0.9) | 73 (1.9) |
| Nepal | 4,188 | 78 (1.9) | 3.02 (0.80) | 2,179 (52.0) | 1,847 (44.1) | 2.68 (1.39) | 1501 (35.8) | 429 (10.2) | 1,112 (26.6) |
| Pakistan | 67,612 | 5,579 (8.3) | 3.02 (0.81) | 34,950 (51.7) | 51,030 (75.5) | 2.66 (1.38) | 28,674 (42.4) | 5,448 (8.1) | 17,859 (26.4) |
| Palestine | 3,707 | 88 (2.4) | 2.95 (0.81) | 1,924 (51.9) | 1,571 (42.4) | 3.09 (1.38) | 270 (7.3) | 42 (1.1) | 65 (1.8) |
| Samoa | 1,571 | 115 (7.3) | 3.00 (0.82) | 824 (52.5) | 1,166 (74.2) | 2.94 (1.41) | 115 (7.3) | 29 (1.8) | 51 (3.2) |
| Sao Tome and Principe | 1,162 | 70 (6.0) | 3.00 (0.83) | 595 (51.2) | 473 (40.7) | 2.85 (1.39) | 154 (13.3) | 37 (3.2) | 61 (5.2) |
| Sierra Leone | 7,088 | 514 (7.3) | 3.01 (0.82) | 3,515 (49.6) | 5,026 (70.9) | 2.54 (1.31) | 2,251 (31.8) | 220 (3.1) | 808 (11.4) |
| Suriname | 2,709 | 92 (3.4) | 2.98 (0.81) | 1,422 (52.5) | 904 (33.4) | 2.66 (1.40) | 173 (6.4) | 110 (4.1) | 152 (5.6) |
| Togo | 2,973 | 226 (7.6) | 3.01 (0.82) | 1,537 (51.7) | 2,061 (69.3) | 2.76 (1.39) | 824 (27.7) | 133 (4.5) | 465 (15.6) |
| Tunisia | 2,171 | 79 (3.6) | 3.06 (0.81) | 1,098 (50.6) | 818 (37.7) | 2.87 (1.37) | 176 (8.1) | 33 (1.5) | 22 (1.0) |
| Zimbabwe | 3,757 | 149 (4.0) | 3.01 (0.82) | 1,849 (49.2) | 2,639 (70.2) | 2.87 (1.42) | 915 (24.4) | 77 (2.0) | 348 (9.3) |

### Underweight

Across all countries, children with disabilities were more likely to be underweight compared to children without disabilities (Table 2: aRR: 1.33, 95% C.I. 1.17 – 1.51). While many samples had small numbers and wide confidence intervals, there was evidence children with disabilities were significantly more likely to be underweight in 13 countries. However, in Ghana (aRR: 0.65, 95% C.I.: 0.45 – 0.95) and Suriname (aRR: 0.12, 95% C.I.: 0.02 – 0.88), children with disabilities were less likely to be underweight than children without disabilities.

**Table 2:** Unadjusted and Adjusted Risk Ratios for Stunting, Wasting, and Underweight for Children with Disabilities Compared to Children without Disabilities.

|  | Underweight |  | Wasted |  | Stunted |  |
| --- | --- | --- | --- | --- | --- | --- |
|  | RR<br>[95% C.I.] | aRR<br>[95% C.I.] | RR<br>[95% C.I.] | aRR<br>[95% C.I.] | RR<br>[95% C.I.] | aRR<br>[95% C.I.] |
| All children | 1.40<br>[1.22, 1.60] | 1.33<br>[1.17, 1.51] | 1.32<br>[1.18, 1.48] | 1.28<br>[1.18, 1.39] | 1.24<br>[1.18, 1.32] | 1.16<br>[1.11, 1.20] |
| Girls | 1.48<br>[1.25, 1.74] | 1.40<br>[1.20, 1.63] | 1.51<br>[1.37, 1.68] | 1.47<br>[1.32, 1.63] | 1.28<br>[1.18, 1.39] | 1.20<br>[1.12, 1.28] |
| Boys | 1.30<br>[1.18, 1.43] | 1.25<br>[1.14, 1.37] | 1.28<br>[1.05, 1.56] | 1.28<br>[1.04, 1.58] | 1.21<br>[1.14, 1.28] | 1.14<br>[1.10, 1.17] |

In terms of sex differences, both girls (aRR: 1.40, 95% C.I.: 1.20 – 1.63) and boys (aRR: 1.30, 95% C.I.: 1.18 – 1.43) with disabilities were significantly more likely to be underweight than girls and boys without disabilities, respectively. For girls, there was significant evidence from nine countries that girls with disabilities were more likely to be underweight than girls without disabilities, while there was no evidence that girls with disabilities were less to be underweight than girls without disabilities in any country. Boys were also more likely to be underweight in eleven countries, though there was evidence from Ghana that boys with disabilities were less likely to be underweight than boys without disabilities (aRR: 0.55, 95% C.I.: 0.32 – 0.95).

### Wasted

Children with disabilities were significantly more likely to be wasted than children without disabilities (aRR: 1.28, 95% C.I.: 1.18 – 1.39) across all countries. Children with disabilities were at greater risk of being wasted in eight countries, although the small number of children with disabilities with wasting resulted in wide confidence intervals for all countries. There was no evidence to suggest that children with disabilities were less likely to be wasted than children without disabilities in any country.

Girls with disabilities were significantly more likely to be wasted than girls without disabilities (aRR: 1.47, 95% C.I.: 1.32 – 1.63) globally. Most countries showed no differences between girls with and without disabilities, except for Chad, Madagascar, Malawi, Mongolia, Pakistan, and State of Palestine, where significantly higher rates of wasting were observed. Among boys, those with disabilities had significantly higher likelihood of being wasted than those without (aRR: 1.28, 95% C.I.: 1.04 -1.58). In four countries, boys with disabilities were significantly more likely to be wasted than boys without disabilities, whilst none of the countries indicated evidence that boys with disabilities were less likely to be wasted than boys without disabilities.

### Stunted

Children with disabilities were significantly more likely to be stunted than children without disabilities (aRR: 1.16, 95% C.I.: 1.11 – 1.20). In 14 countries, children with disabilities had a higher likelihood of stunting, while no countries showed evidence of children with disabilities being less likely to be stunted. Both girls and boys with disabilities had significantly higher rates of stunting compared to their counterparts without disabilities (girls: aRR: 1.20, 95% C.I. 1.12 - 1.28, boys: prevalence aRR: 1.14, 95% C.I. 1.10 – 1.17). For each sex, there was no evidence the children with disabilities had lower prevalence of stunting than children without disabilities. However, most countries had small sample sizes and wide confidence intervals.

## Discussion

Using comparable data from 30 countries, we found that young children with disabilities are significantly more likely to be stunted, wasted, and underweight than children without disabilities. In sex-disaggregated analyses, both boys and girls are also significantly more likely to be malnourished than boys and girls without disabilities, respectively. The findings presented here have profound implications for meeting the lifelong impacts of malnutrition in childhood, as programmes will need to work to address the disproportionate prevalence of malnutrition on children with disabilities. These findings also highlight that achieving SDG 2 and global child mortality reduction targets will be impossible without a sufficient focus on disability-inclusion.

Our study adds to the body of evidence that has shown higher prevalence and adverse impacts of nutritional disorders in young children with disabilities compared to those without disabilites.^4^ A 2017 systematic review of 17 studies found that children with disabilities in LMICs were nearly three times more likely to be underweight and nearly two times more likely to be wasted or stunted compared to children without disabilities.^4^ Other studies have also produced evidence for specific impairments and/or geographic locations. For example, a systematic review of malnutrition among children and adolescents with cerebral palsy in Arab-speaking countries found that children with cerebral palsy had substantially higher prevalence of malnutrition.^11^ Prior evidence from Malawi has also showed that children with disabilities were more likely to have adverse outcomes from severe acute malnutrition than children without disabilities.^5^

Our findings have a range of policy and programmatic implications. Firstly, while there has been increasing focus on addressing various social inequities in malnutrition programmes, these have been insufficient with regards to disability.^26^ Various barriers exist for caregivers of children with disabilities to access health and nutritional services,^22^ ^36^ as these data provide further evidence that urgent action is needed to close these gaps. A health systems approach can play a crucial role in addressing these differences for children with disabilities. For example, given the lack of disability-specific guidelines on nutrition programming and invisibility in mainstream nutrition programmes,^26^ governments, international organisations, donors, and NGOs alike can improve how children with disabilities are included in nutrition policies, guidelines, and programmes. In terms of health financing, it is essential key stakeholders develop specific programmes and budget lines to target children with disabilities. Identifying children with disabilities within the primary care system and referring those at risk of malnutrition to care would strengthen coordination between primary care and more specialised services and rehabilitation. Nurses, midwives, skilled birth attendants, and community health workers need to receive training to recognise children with disabilities and nutritional deficiencies, offer precise parental education regarding disabilities, which can help diminish stigma, misinformation,^37^ and potential risks of abuse or neglect for the child, and appropriately refer these children to the required services. Given the higher prevalence of malnutrition outcomes for girls, there is also evidence to suggest a gender-sensitive approach is needed.^36^

Additionally, stakeholders can co-create curricula and programmes for parents of children with disabilities to address some of the stigma and cultural attitudes surrounding feeding and health practices for children with disabilities.^36^ By building awareness and providing financial support and incentives to improve nutrition, parents of children with disabilities can be supported to improve awareness, feeding practices, and outcomes. Prior research has shown that these interventions may be promising to support these parents, and so further expansion of this may be beneficial.^38^ Furthermore, more training for health workers is needed to support identification of children with disabilities, tackle stigma towards children with disabilites,^39^ ^40^ as well as those at risk of malnutrition. Upskilling health workers on disability awareness, addressing stigma, and improving knowledge on malnutrition will help provide earlier intervention and greater support to children with disabilities experiencing malnutrition. However, recent mapping to understand key research gaps for children with disabilities suggests more research is needed to understand the interventions that can help close these inequities for disabled children.^41^

It is crucial for future nutrition policy and programming, maternal and child health, and disability policy to acknowledge and address the connection between malnutrition and disability. This work should be twin tracked to ensure children with disabilities are reached in mainstream efforts, but also ensure that the specific needs of children with disabilities are included. For example, children with disabilities may need to have tailored programs because of additional and specific feeding difficulties (i.e., children with Autism may have difficulty tolerating different food textures)^42^ ^43^ and because of the specific exclusions this population faces (i.e., exclusion from education mean exclude children with disabilities are not included in school-based nutrition programmes).^44^ By doing so, existing challenges can be transformed into opportunities to benefit both areas of healthcare, requiring adequate resources and effective action planning. Including children with disabilities in nutrition services and considering their specific needs will contribute to inclusive and equitable access to nutrition as a fundamental human right.^7^

Finally, all malnutrition programmes should collect disability data to understand how they are reaching children with disabilities, as well as the outcomes for this population. This is particularly important to examine through the lens of different impairments to see if further, targeted interventions are required. Through this system-level approach, it will be possible to ensure that these inequities for children with disabilities are addressed in the global efforts to end all forms of malnutrition by 2030.

### Strengths and Limitations

This is the largest study to date to examine disability and sex-based inequities in key malnutrition outcomes for nearly 230,000 children in household surveys across 30 low- and middle-income countries. The large-scale, high-quality, and internationally comparable UNICEF-supported MICS data provide strong evidence for these inequities and should be used as motivation to address these inequities. However, this analysis also has several limitations.

First, the small numbers of children with certain outcomes means that much of the sex-disaggregated data had small numbers and wide confidence intervals, limiting our ability to draw conclusions about the intersectional barriers children with disabilities may experience. Secondly, the overlap of the Washington Group Questions and the outcome of interest hampers our ability to look at younger children or other important covariates (i.e., breastfeeding) that may impact nutrition outcomes. Finally, the MICS anthropomorphic measurement manual does not mention disability, meaning the growth standards and measurements may not capture all children with disabilities (i.e., a child with short stature is not captured as stunted because it does not use expected height, rather than actual height). Therefore, these results likely underestimate the burden of nutritional disorders amongst children with disabilities.

## Conclusion

Children with disabilities are unacceptably overrepresented in all three key malnutrition indicators—stunting, wasting, and underweight. These relative inequities are not due to impairment alone and need to be urgently addressed in order to reach the SDG targets. Concerted efforts to improve disability in nutrition programmes and throughout the health system is urgently needed. Without a focus on disability, we risk perpetuating inequities in malnutrition and related morality—an unacceptable violation of children with disabilities’ human right to health.

## Public and Patient Involvement

It was not appropriate or possible to involve patients or the public in the design, conduct, reporting, or dissemination of this research as we have used secondary, anonymized data from multiple countries.

## Data Availability

Raw data used in this analysis are available from the Multiple Indicator Cluster Survey website.

https://mics.unicef.org/surveys

---

**Supplementary Figure 1:**
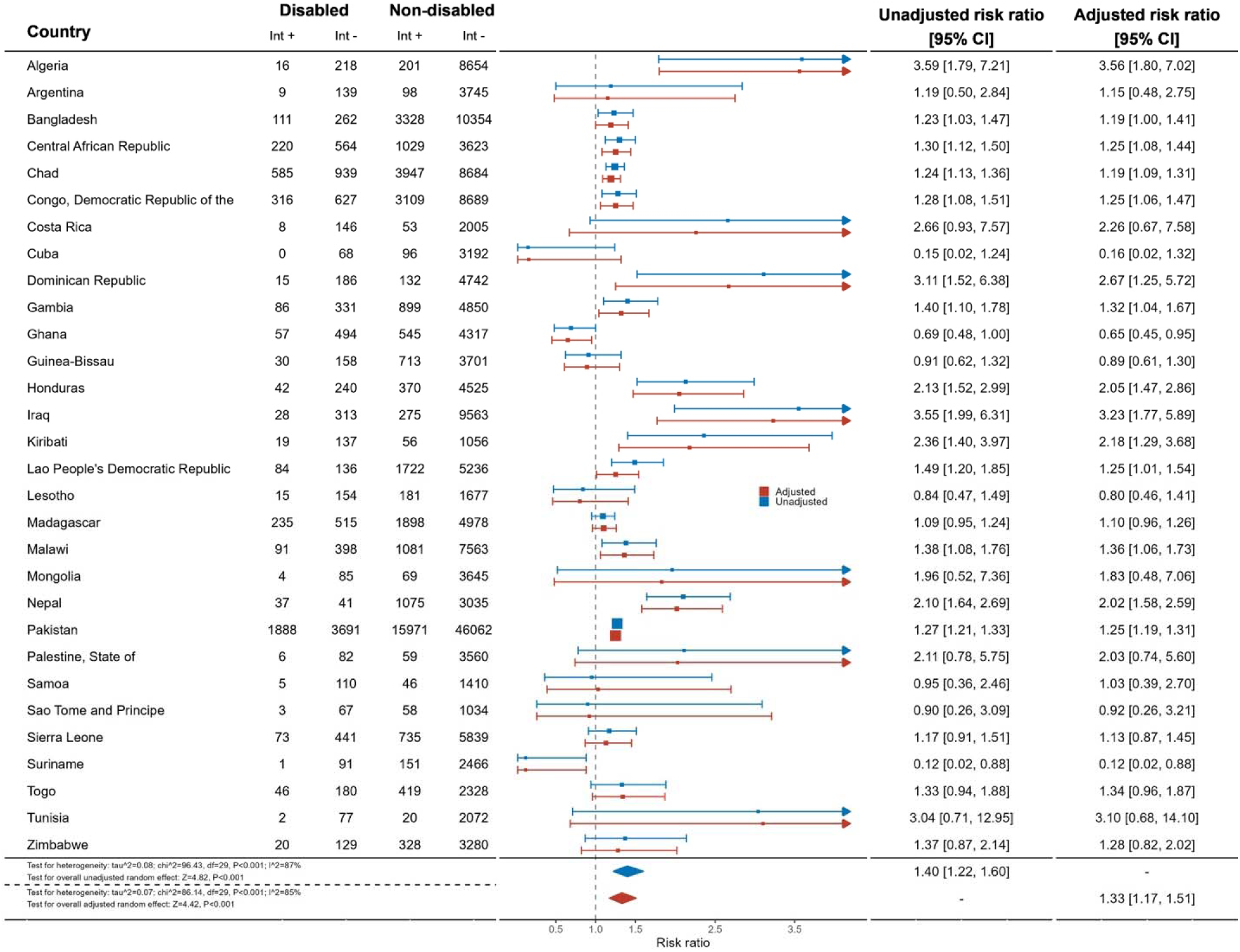
Meta-analysis comparing prevalence of underweight in children with and without disabilities

**Supplementary Figure 2:**
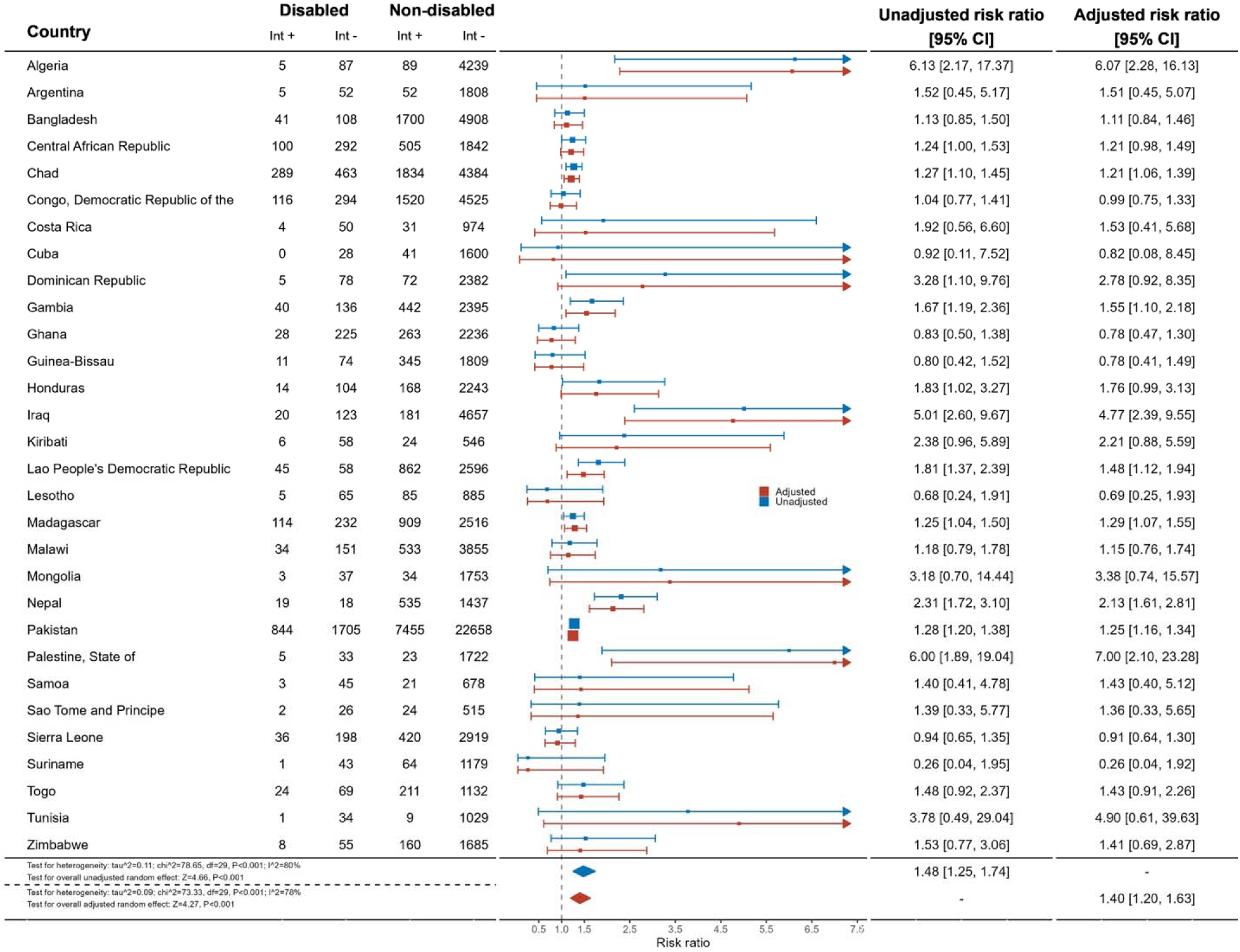
Meta-analysis comparing prevalence of underweight in girls with and without disabilities

**Supplementary Figure 3:**
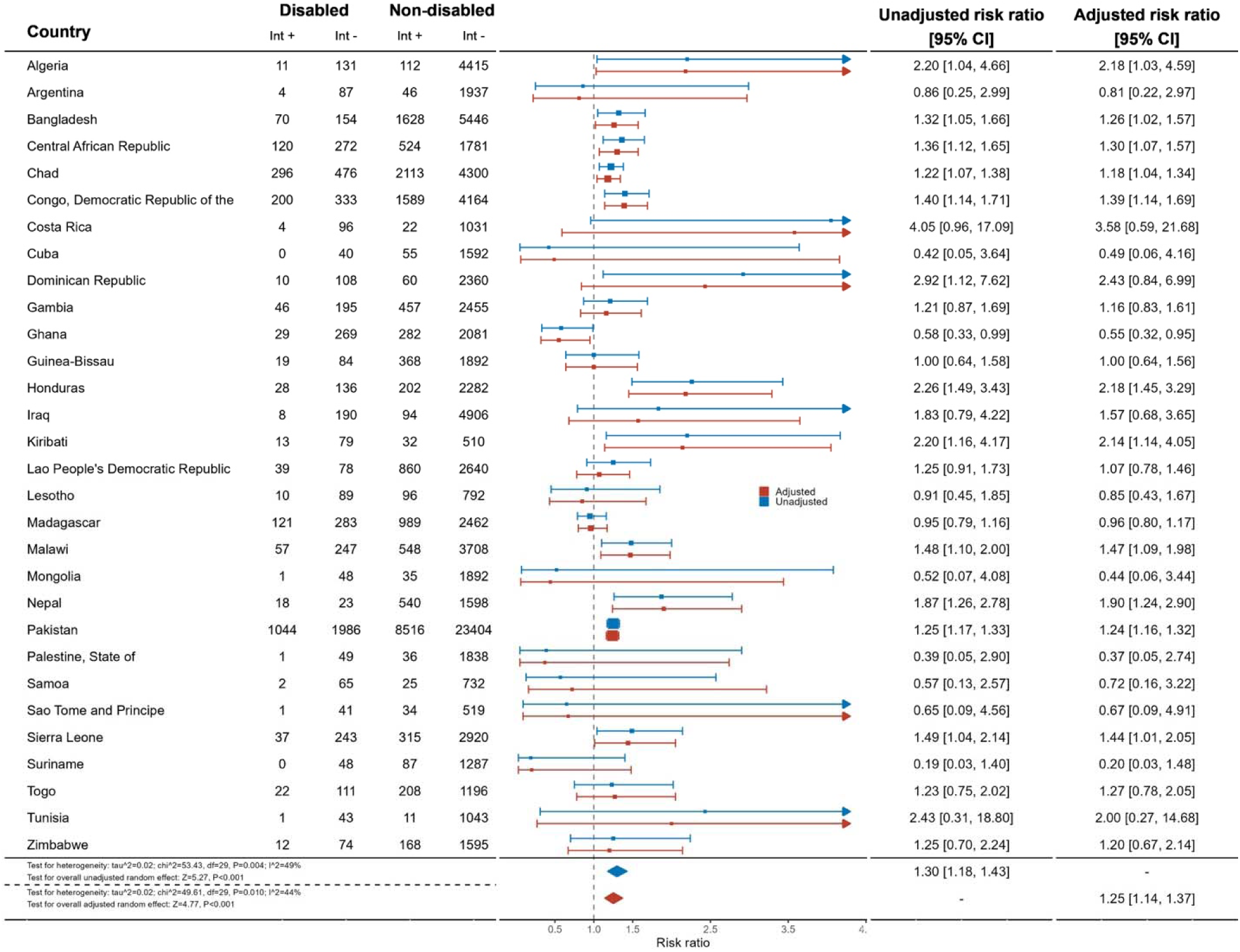
Meta-analysis comparing prevalence of underweight in boys with and without disabilities

**Supplementary Figure 4:**
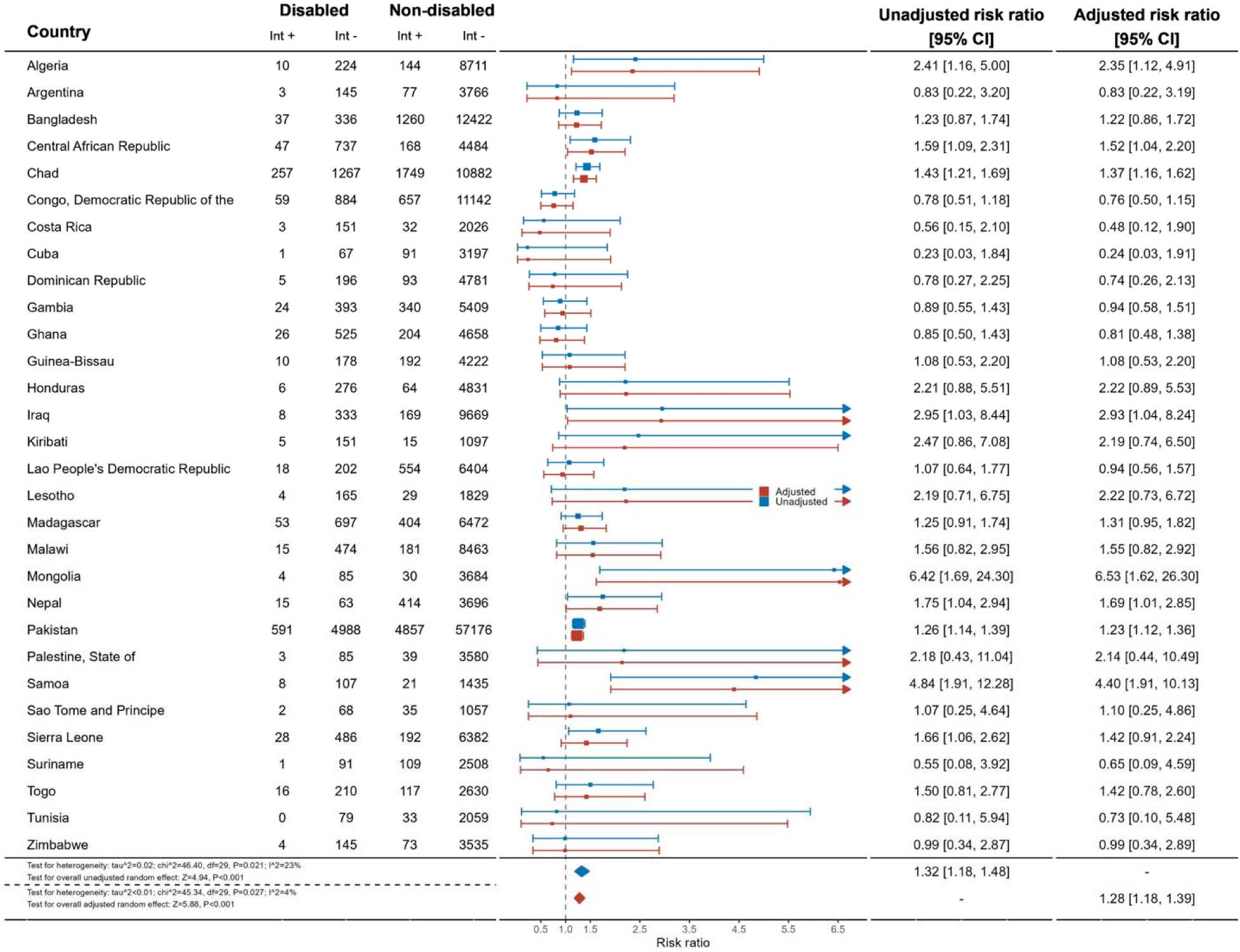
Meta-analysis comparing prevalence of wasting in children with and without disabilities

**Supplementary Figure 5:**
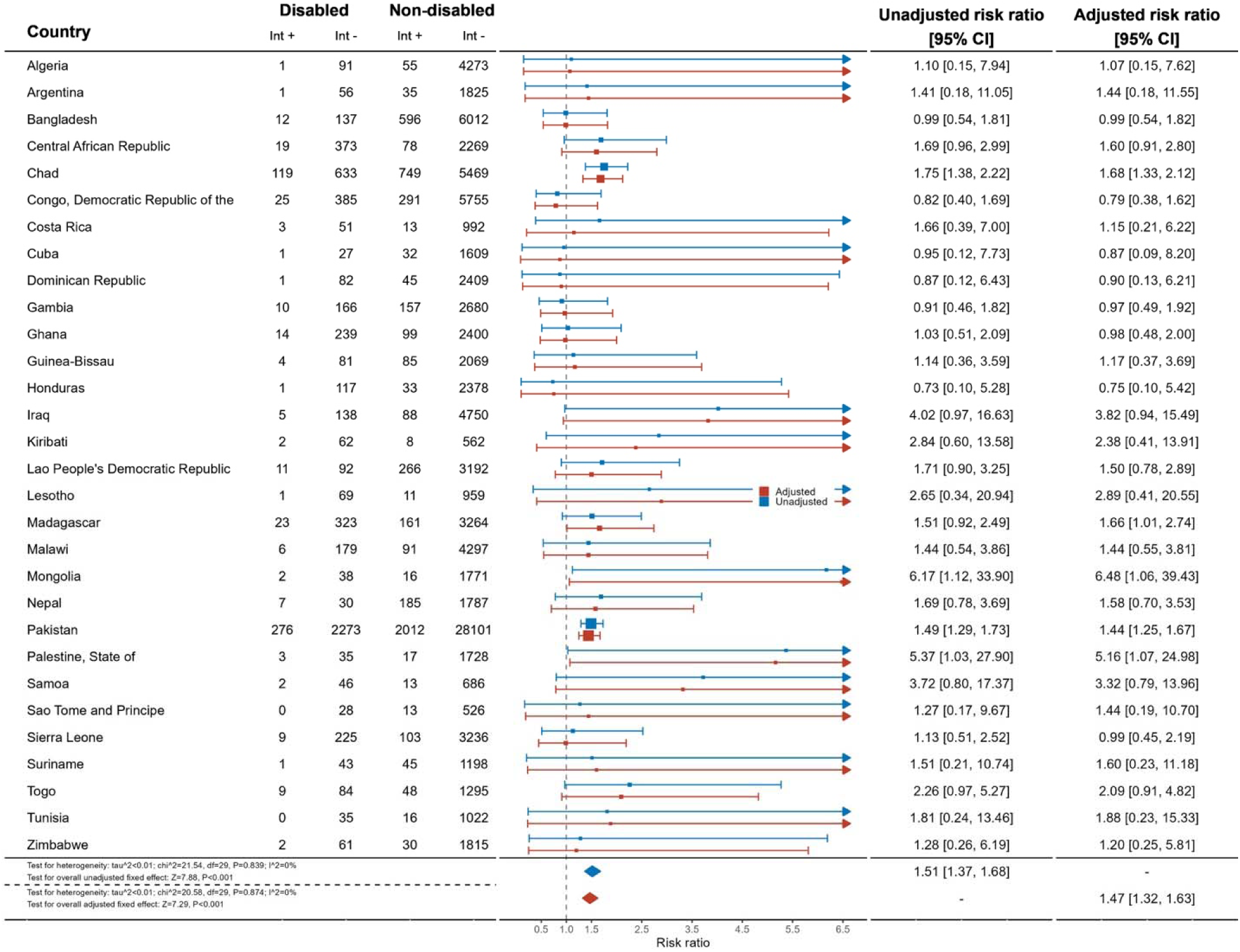
Meta-analysis comparing prevalence of wasting in girls with and without disabilities

**Supplementary Figure 6:**
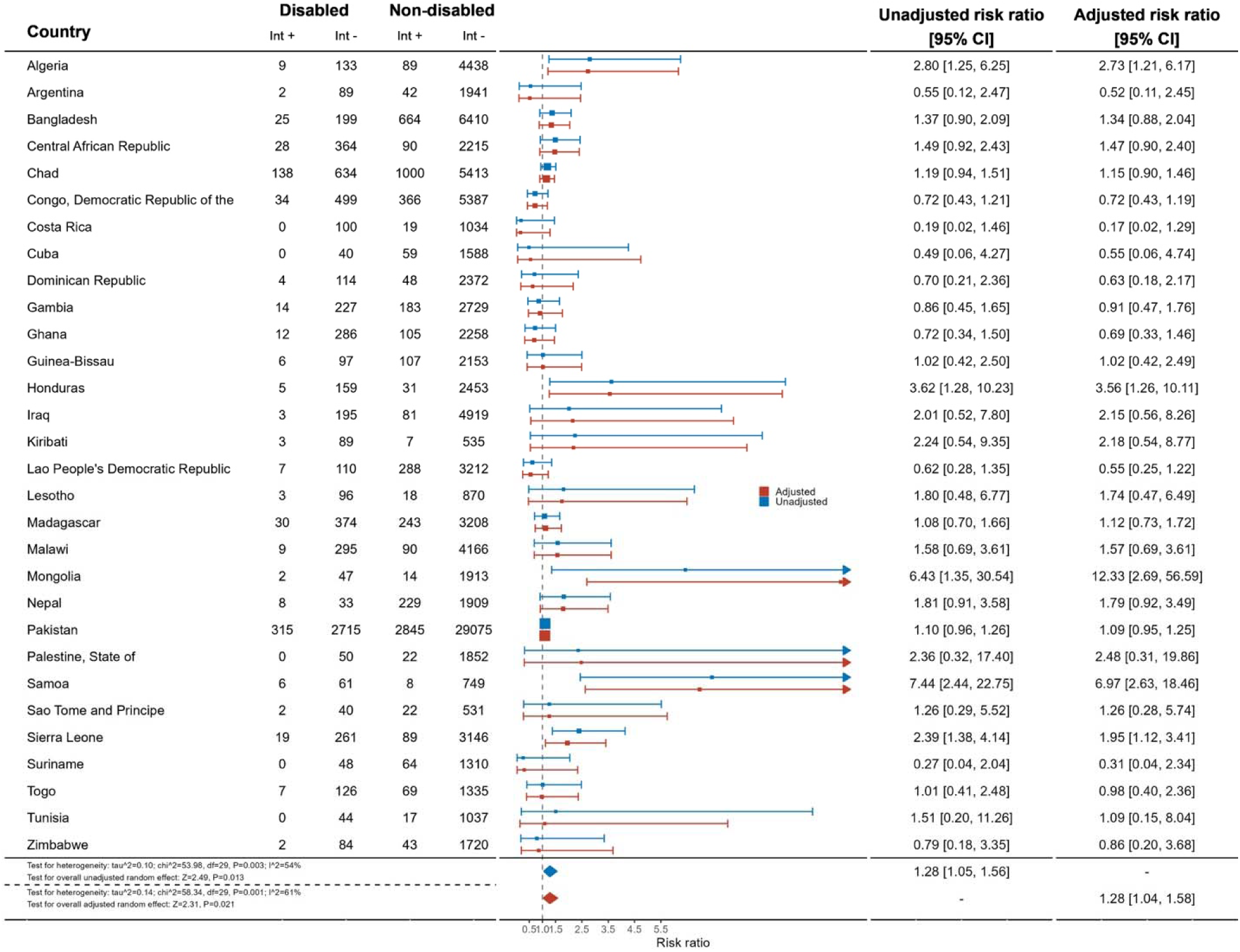
Meta-analysis comparing prevalence of wasting in boys with and without disabilities

**Supplementary Figure 7:**
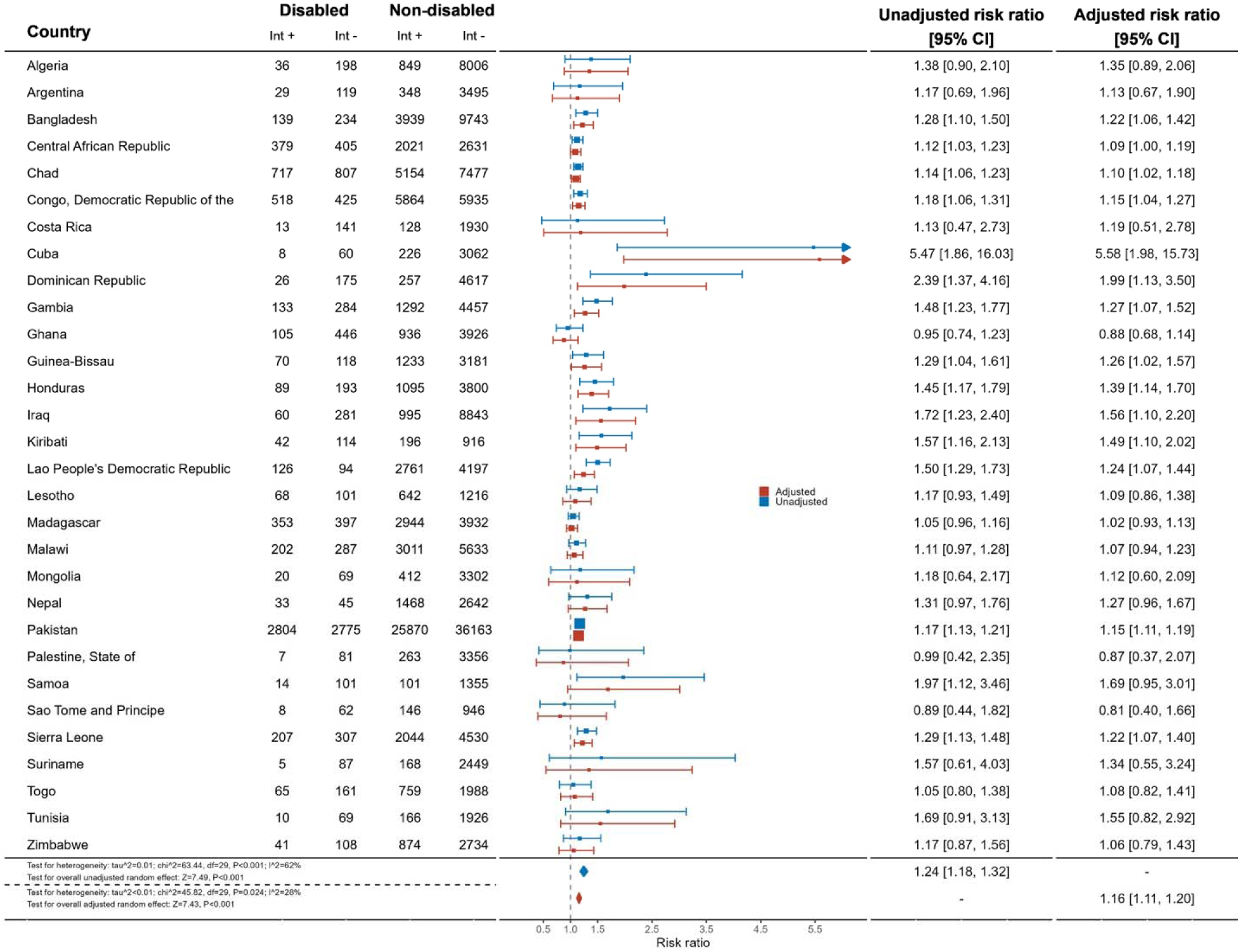
Meta-analysis comparing prevalence of stunting in children with and without disabilities

**Supplementary Figure 8:**
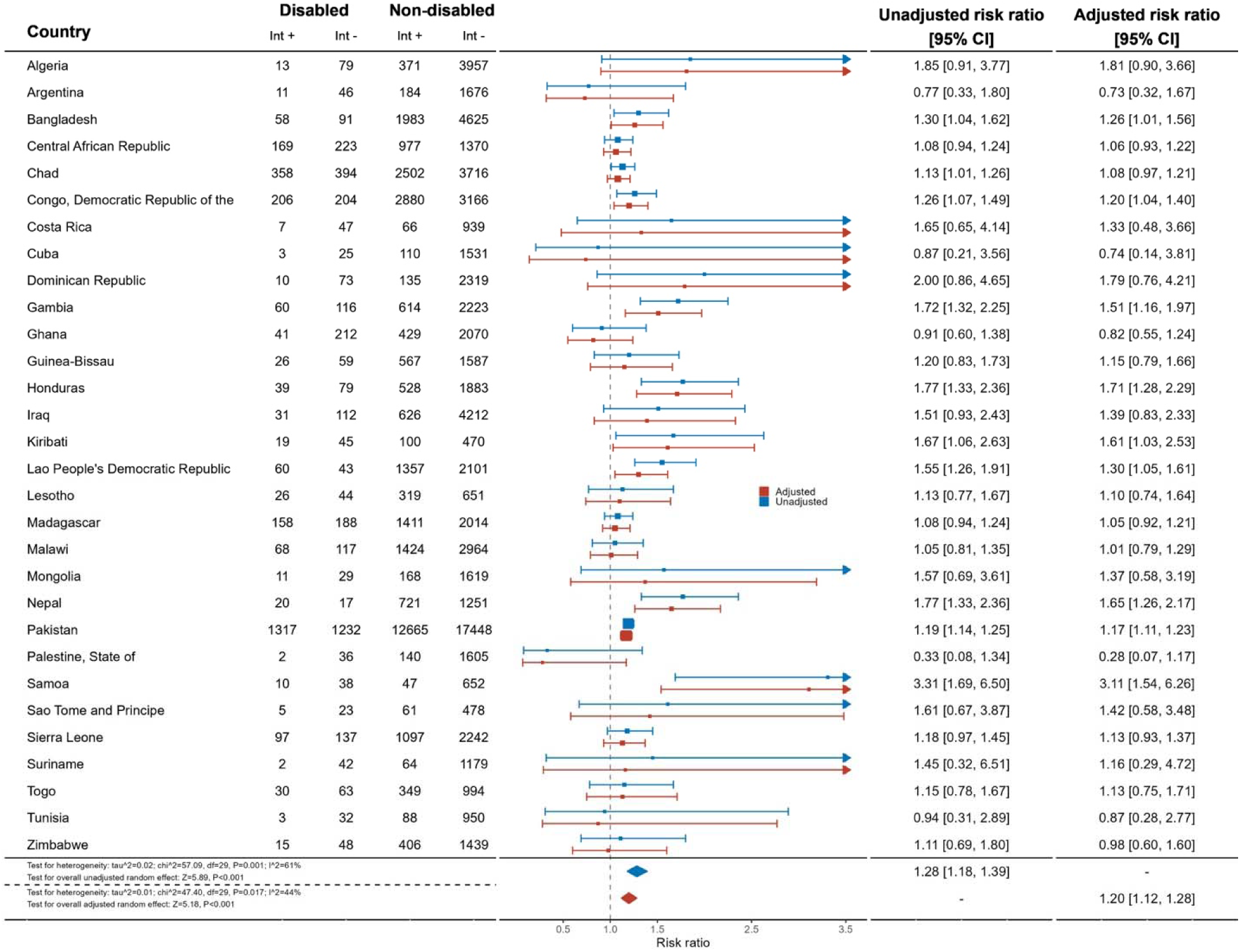
Meta-analysis comparing prevalence of stunting in girls with and without disabilities

**Supplementary Figure 9:**
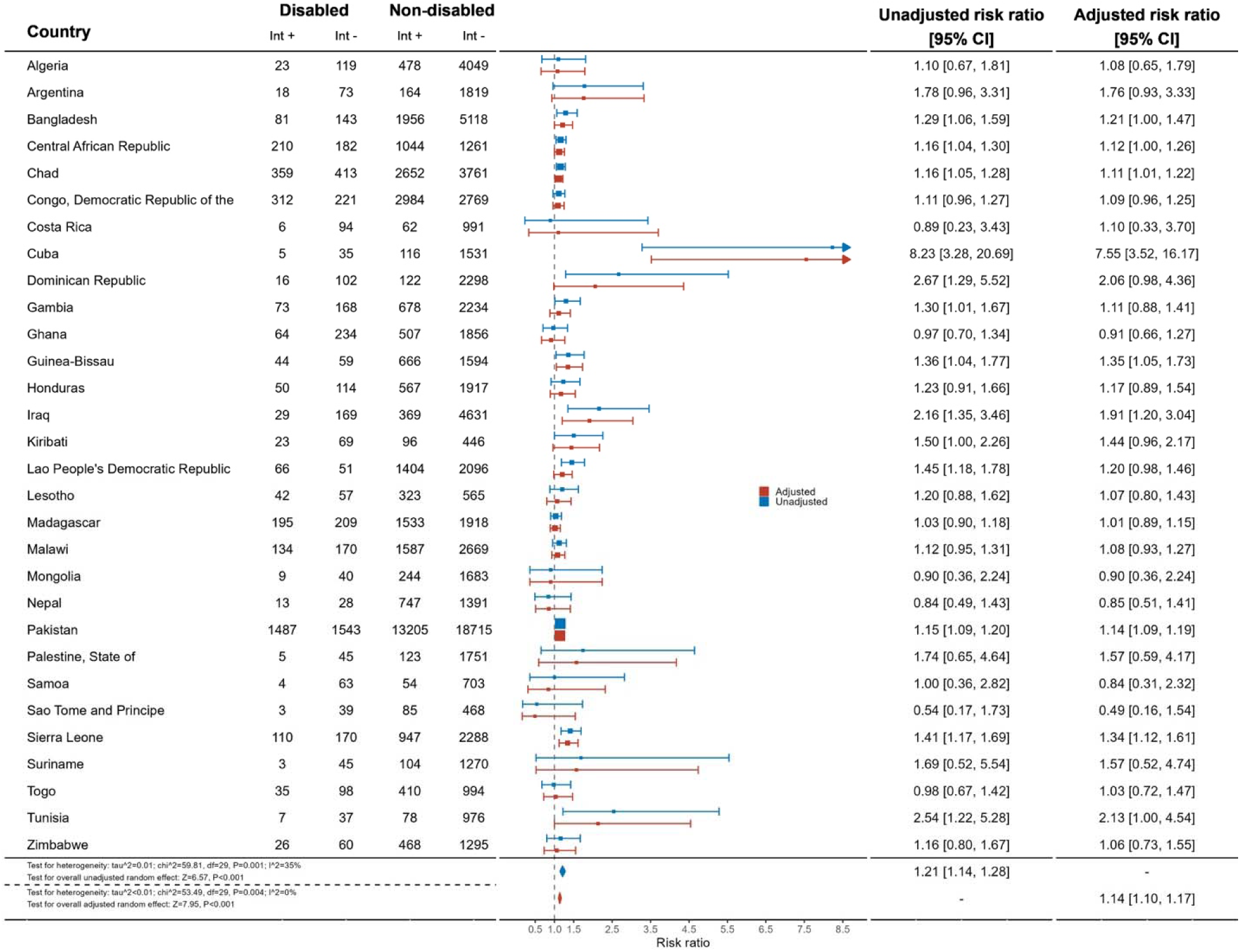
Meta-analysis comparing prevalence of stunting in boys with and without disabilities

## References

1. Kambale RM, Francisca IN. Optimising the management of acute malnutrition. The Lancet Global Health 2022;10(4):e453–e54. doi: 10.1016/S2214-109X(22)00087-0

2. Bhutta ZA, Berkley JA, Bandsma RHJ, et al. Severe childhood malnutrition. Nature Reviews Disease Primers 2017;3(1):17067. doi: 10.1038/nrdp.2017.67

3. UNICEF. Seen, Counted, Included: Using data to shed light on the well-being of children with disabilities. New York: United Nations Children’s Fund, 2021.

4. Hume-Nixon M, Kuper H. The association between malnutrition and childhood disability in low- and middle-income countries: systematic review and meta-analysis of observational studies. Tropical Medicine & International Health 2018;23(11):1158–75. doi: 10.1111/tmi.13139 [published Online First: 20180910]

5. Kerac M, Chagaluka G, Kett M, et al. Impact of disability on survival from severe acute malnutrition in a developing country setting - a longitudinal cohort study. Archives of Disease in Childhood 2012;97(Suppl 1):A43–A44. doi: 10.1136/archdischild-2012-301885.107

6. Jacobs AE. How Body Mass Index Compromises Care of Patients With Disabilities. AMA J Ethics 2023;25(7):E545–49.

7. Groce N, Challenger E, Berman-Bieler R, et al. Malnutrition and disability: unexplored opportunities for collaboration. Paediatr Int Child Health 2014;34(4):308–14. doi: 10.1179/2046905514y.0000000156 [published Online First: 20141013]

8. Kasajja M, Nabiwemba E, Wamani H, et al. Prevalence and factors associated with stunting among children aged 6-59 months in Kabale district, Uganda. BMC Nutr 2022;8(1):79. doi: 10.1186/s40795-022-00578-9 [published Online First: 20220815]

9. World Health Organization. Essential nutrition actions: improving maternal, newborn, infant and young child health and nutrition. 2013

10. Lelijveld N, Groce N, Patel S, et al. Long-term outcomes for children with disability and severe acute malnutrition in Malawi. BMJ Glob Health 2020;5(10):e002613. doi: 10.1136/bmjgh-2020-002613

11. Mushta SM, Jahan I, Sultana R, et al. Burden of Malnutrition among Children and Adolescents with Cerebral Palsy in Arabic-Speaking Countries: A Systematic Review and Meta-Analysis. Nutrients 2021;13(9) doi: 10.3390/nu13093199 [published Online First: 20210915]

12. Boudokhane S, Migaou H, Kalai A, et al. Feeding problems and malnutrition associated factors in a North African sample of multidisabled children with cerebral palsy. Res Dev Disabil 2021;118:104084. doi: 10.1016/j.ridd.2021.104084 [published Online First: 20210917]

13. Skrzypek M, Koch W, Goral K, et al. Analysis of the Diet Quality and Nutritional State of Children, Youth and Young Adults with an Intellectual Disability: A Multiple Case Study. Preliminary Polish Results. Nutrients 2021;13(9) doi: 10.3390/nu13093058 [published Online First: 20210831]

14. Batra A, Marino LV, Beattie RM. Feeding children with neurodisability: challenges and practicalities. Arch Dis Child 2022;107(11):967–72. doi: 10.1136/archdischild-2021-322102 [published Online First: 20220201]

15. Amadu I, Seidu AA, Duku E, et al. Risk factors associated with the coexistence of stunting, underweight, and wasting in children under 5 from 31 sub-Saharan African countries. BMJ Open 2021;11(12):e052267. doi: 10.1136/bmjopen-2021-052267 [published Online First: 20211220]

16. Banks LM, Kuper H, Polack S. Poverty and disability in low-and middle-income countries: A systematic review. PLOS ONE 2017;12(12):e0189996.

17. Emerson E, Llewellyn G. Identifying children at risk of intellectual disability in UNICEF’s multiple indicator cluster surveys: Cross-sectional survey. Disability and Health Journal 2021;14(1):100986. doi: 10.1016/j.dhjo.2020.100986

18. White S, Kuper H, Itimu-Phiri A, et al. A Qualitative Study of Barriers to Accessing Water, Sanitation and Hygiene for Disabled People in Malawi. PLOS ONE 2016;11(5):e0155043. doi: 10.1371/journal.pone.0155043

19. Kuhlthau KA, Perrin JM. Child Health Status and Parental Employment. Archives of Pediatrics & Adolescent Medicine 2001;155(12):1346–50. doi: 10.1001/archpedi.155.12.1346

20. World Bank, World Health Organization. World Report on Disability, 2011:350.

21. Rotenberg S, Davey C, McFadden E. Association between disability status and health care utilisation for common childhood illnesses in 10 countries in sub-Saharan Africa: a cross-sectional study in the Multiple Indicator Cluster Survey. eClinicalMedicine 2023;57 doi: 10.1016/j.eclinm.2023.101870

22. The Missing Billion Initiative, Clinton Health Access Initative. Reimagining Health Systems That Expect, Accept and Connect 1 Billion People with Disabilities, 2022:30.

23. Winskill P, Hogan AB, Thwing J, et al. Health inequities and clustering of fever, acute respiratory infection, diarrhoea and wasting in children under five in low- and middle-income countries: a Demographic and Health Surveys analysis. BMC Med 2021;19(1):144. doi: 10.1186/s12916-021-02018-0 [published Online First: 20210624]

24. UNICEF. Child Malnutrition New York2023 [accessed August 1 2023.

25. Olusanya BO, Wright SM, Nair MKC, et al. Global Burden of Childhood Epilepsy, Intellectual Disability, and Sensory Impairments. Pediatrics 2020;146(1) doi: 10.1542/peds.2019-2623 [published Online First: 20200617]

26. Engl M, Binns P, Trehan I, et al. Children living with disabilities are neglected in severe malnutrition protocols: a guideline review. Arch Dis Child 2022;107(7):637–43. doi: 10.1136/archdischild-2021-323303 [published Online First: 20220204]

27. The Global Goals. 2: Zero Hunger 2: Zero Hunger [Available from: https://www.globalgoals.org/goals/2-zero-hunger/?gclid=Cj0KCQjw4s-kBhDqARIsAN-ipH1zJCXgv3Hr0m0ow36HpMgEvMaDetciK4BYFWEUukQb1YqHejZNwjMaAvSnEALw_wc B accessed June 22 2023.

28. Hashemi G, Kuper H, Wickenden M. SDGs, inclusive health and the path to universal health coverage. Disability and the global south. Disability and the Global South 2017;4(1):1088–111.

29. UNICEF. MICS Survey Database. 2020

30. Khan S, Hancioglu A. Multiple Indicator Cluster Surveys: Delivering Robust Data on Children and Women across the Globe. Studies in Family Planning 2019;50(3):279–86. doi: 10.1111/sifp.12103

31. Word Health Organization. WHO Child Growth Standards Geneva2006 [Available from: https://www.who.int/tools/child-growth-standards accessed 21 June 2023.

32. World Health Organization. WHO child growth standards : training course on child growth assessment. Geneva: World Health Organization, 2008.

33. Multiple Indicator Cluster Survey Team. Review of Options for Reporting Water, Sanitation, and Hygeine Coverage by Wealth Quintile. MICS Methodological Papers. New York: UNICEF, 2016:141.

34. Zou G. A Modified Poisson Regression Approach to Prospective Studies with Binary Data. American Journal of Epidemiology 2004;159(7):702–06. doi: 10.1093/aje/kwh090

35. Lumley T. Survey: analysis of complex survey samples. R package version 4.0., 2020.

36. Holden J, Corby N. Disability and nutrition programming: evidence and learning In: Report UADIH, ed. London: UKAID, 2019.

37. World Health Organization. Survive and thrive: transforming care for every small and sick newborn. Geneva: World Health Organization 2019:x, 150 p.

38. Zuurmond M, O’Banion D, Gladstone M, et al. Evaluating the impact of a community-based parent training programme for children with cerebral palsy in Ghana. PLOS ONE 2018;13(9):e0202096. doi: 10.1371/journal.pone.0202096 [published Online First: 20180904]

39. Shakespeare T, Iezzoni LI, Groce NE. Disability and the training of health professionals. The Lancet 2009;374(9704):1815-16. doi: 10.1016/S0140-6736(09)62050-X

40. Rotenberg S, Gatta DR, Wahedi A, et al. Disability Training for Health Workers: A Global Evidence Synthesis. Disability and Health Journal 2022:101260.

41. Thota A, Mogo E, Igbelina D, et al. Inclusion Matters: Inclusive Interventions for Children with Disabilities – An evidence and gap map from low- and middle-income countries. In: UNICEF Office of Research - Innocenti Florence, ed. Innocenti Research Report, 2022.

42. Manikandan B, Gloria J. K, Samuel R, et al. Feeding Difficulties Among Children With Special Needs: A Cross-Sectional Study From India. OTJR: Occupational Therapy Journal of Research 2022;0(0):15394492221130971. doi: 10.1177/15394492221130971

43. Andrew MJ, Sullivan PB. Feeding difficulties in disabled children. Paediatrics and Child Health 2010;20(7):321–26. doi: 10.1016/j.paed.2010.02.005

44. Meresman S, Drake L. Are School Feeding Programs Prepared to Be Inclusive of Children with Disabilities? Frontiers in Public Health 2016;4 doi: 10.3389/fpubh.2016.00045

